# Extended Reality for Neuraxial Anesthesia and Pain Procedures: A Scoping Review

**DOI:** 10.1101/2024.01.29.24301926

**Authors:** James S. Cho, Devaunsh M. Thaker, Rohan Jotwani, David Hao

## Abstract

**Background:** Extended reality technology, encompassing augmented reality, mixed reality, and virtual reality, has the potential to enhance the teaching and performance of neuraxial procedures. The diverse applications of extended reality include immersive simulations and novel modes of procedural navigation.

**Objectives:** This scoping review aims to explore the preclinical, clinical, and educational applications of extended reality for neuraxial procedures while suggesting directions for future research.

**Evidence review:** A systematic search was conducted across PubMed, Embase, Web of Science, Cochrane Central Register of Controlled Trials, and Google Scholar until December 2023. Additional sources were identified via citation searching of relevant articles. The findings are reported using the Preferred Reporting Items for Systematic Reviews and Meta-Analyses extension for Scoping Reviews (PRISMA-ScR).

**Findings:** 41 studies, including three pending clinical trials were included. The majority of included studies were published after 2015. Extended reality technology was applied in diverse ways for teaching, simulation, and navigation, but only four of the completed studies described clinical use. For the display of visuals, computer screens were most commonly used, followed by head-mounted devices, laser projectors, and semi-transparent mirrors.

**Conclusions:** Interest in utilizing extended reality technology for neuraxial procedures is growing. Preliminary work shows promise for this technology in both education and clinical practice, but achieving accurate image registration without disrupting existing workflows remains an ongoing barrier to clinical testing. Additional research is needed to assess the cost-effectiveness and reliability of this technology.

## INTRODUCTION

Extended reality (XR) (encompassing augmented reality [AR] where digital objects overlay onto physical surroundings, mixed reality [MR] where digital and physical objects interact, and virtual reality [VR] where users interact with digital scenes and objects) holds significant potential in medical education and clinical practice.[1–3] XR can be used to create immersive simulations or enhance standard modes of anatomic navigation.[2] Despite these potential benefits, the use of XR within anesthesiology and pain medicine remains limited.[4]

Neuraxial procedures for anesthesia and pain medicine are commonly taught and performed using landmark-based approaches or with the use of ultrasound or fluoroscopy.[5,6] However, each method has limitations related to accuracy, ergonomics, or radiation exposure. The evolution of XR technology, combined with improved access to spatial computing devices, could pave the way for innovative methods of teaching and performing neuraxial procedures. With sufficient development and validation, XR technology may improve the quality of education and patient care. However, the current progress in adopting this technology for neuraxial procedures has not been systematically analyzed.

This scoping review aims to summarize the current applications of XR for neuraxial procedures and to identify barriers to widespread use. The following questions were defined to inform our search strategy:

1. How has extended reality been used for neuraxial procedures in clinical settings?
2. How has extended reality been used for training clinicians to perform neuraxial procedures?
3. What are the benefits of and barriers to adopting XR for teaching or performing neuraxial procedures?
4. What are clinician attitudes towards adopting XR for teaching or performing neuraxial procedures?

## METHODS

A protocol for this scoping review was preregistered on the Open Science Framework on November 5th, 2023.[7] The reporting of this scoping review was guided by the standards of the Preferred Reporting Items for Systematic Reviews and Meta-Analyses extension for Scoping Reviews (PRISMA-ScR).[8]

### Search Strategy and Sources of Evidence

A systematic search of the literature was performed on PubMed, Embase, Web of Science, Cochrane Central Register of Controlled Trials, and Google Scholar. The search query combined ((augmented OR extended OR mixed OR virtual) AND reality) AND (epidural OR peridural OR intrathecal OR spinal OR neuraxial) AND (anesthesia OR pain). No filters were applied. Due to a high volume of irrelevant results on Google Scholar (greater than 200,000 results), we initially screened the first 100 results for relevance, and later expanded to include 20 additional results in the most recent search on December 14th, 2023. Additional references were identified through citation searching of relevant articles. Pending trials were closely monitored, and the most recent trial included was published on January 11th, 2024. The search strategy used for each database is provided in the online supplemental appendix.

### Eligibility Criteria

The PICO (population, intervention, comparison, and outcomes) framework was used to develop the inclusion and exclusion criteria (**table 1**). All primary studies available in English and investigating the use of XR in the performance or teaching of neuraxial procedures were included. Technical reports and conference papers were also considered. No restrictions were imposed on the publication year. Although review articles were ultimately excluded, each related review underwent thorough evaluation, and relevant citations from these reviews were incorporated into the screening pool. Essays and letters describing theoretical applications were excluded.

**Table 1.** Inclusion and exclusion criteria.

|  | Inclusion Criteria | Exclusion Criteria |
| --- | --- | --- |
| Population | Clinicians and trainees performing or learning neuraxial procedures; patients receiving neuraxial procedures | No additional limitations |
| Intervention | Use of extended reality to aid the teaching or performance of neuraxial anesthesia or pain procedures | Use of extended reality by patient (e.g. for anxiolysis during a procedure) |
| Comparison | All comparisons | No limitations |
| Outcomes | All outcomes | No limitations |
| Study Design | Any primary investigations | Reviews<sup>a</sup> and essays or letters describing theoretical applications |
| Years | Any | No limitations |
| Language | English full-text available | English full-text not available |

a: Reviews were excluded, but relevant bibliography was screened and included as appropriate

### Study Selection

After importing all retrieved articles into Covidence (Melbourne, Australia), automatic and manual deduplication was performed. Two authors (JSC and DMT) screened the articles initially by title and abstract, followed by a full-text review. When access to full-text was unavailable, corresponding authors were contacted for access. A third author (DH) was available to provide resolution if discrepancies could not be resolved via discussion.

### Data Extraction

Two authors (JSC and DMT) manually extracted the following data into pre-designed forms on Covidence: title, authors, year of publication, first author country, study design, purpose of XR system, mode of XR, XR display device used, procedure taught or performed, number of participants and description, key outcomes, clinician attitudes regarding adoption of XR, limitations of described system, and suggestions for further research. For this review, AR and MR were grouped into one category, as explained later in the study. Study designs were often not explicitly stated and were manually coded by the authors.

To maximize congruence, we used an iterative approach where the two extractors periodically compared their results (initially after each study and subsequently after every few studies), achieved consensus, and discussed strategies to maximize accuracy. As with screening, a third author (DH) was available to resolve disagreements.

### Synthesis of Results

We grouped the included studies by the purpose for which XR applications were created and summarized the device(s) used to achieve XR, the neuraxial procedure(s) performed, the population studied, and key findings. Participants’ responses to the use of XR as well as notable limitations of the described system were also summarized. Study authors’ suggestions for further research were also tabulated.

## FINDINGS

A PRISMA flowchart illustrating our review is shown in **figure 1**. Our initial search yielded 824 entries, and citation searching led to the addition of 28 additional references. One unindexed article co-written by one of the authors (RJ) was manually included, as it met inclusion criteria. 280 duplicates were removed. After screening of titles and abstracts, 54 articles were selected for full-text retrieval. All 54 full-text articles were successfully retrieved (three after contacting the authors), and 41 studies were ultimately included in this review.References pertaining to the same study were merged.

**Figure 1:**
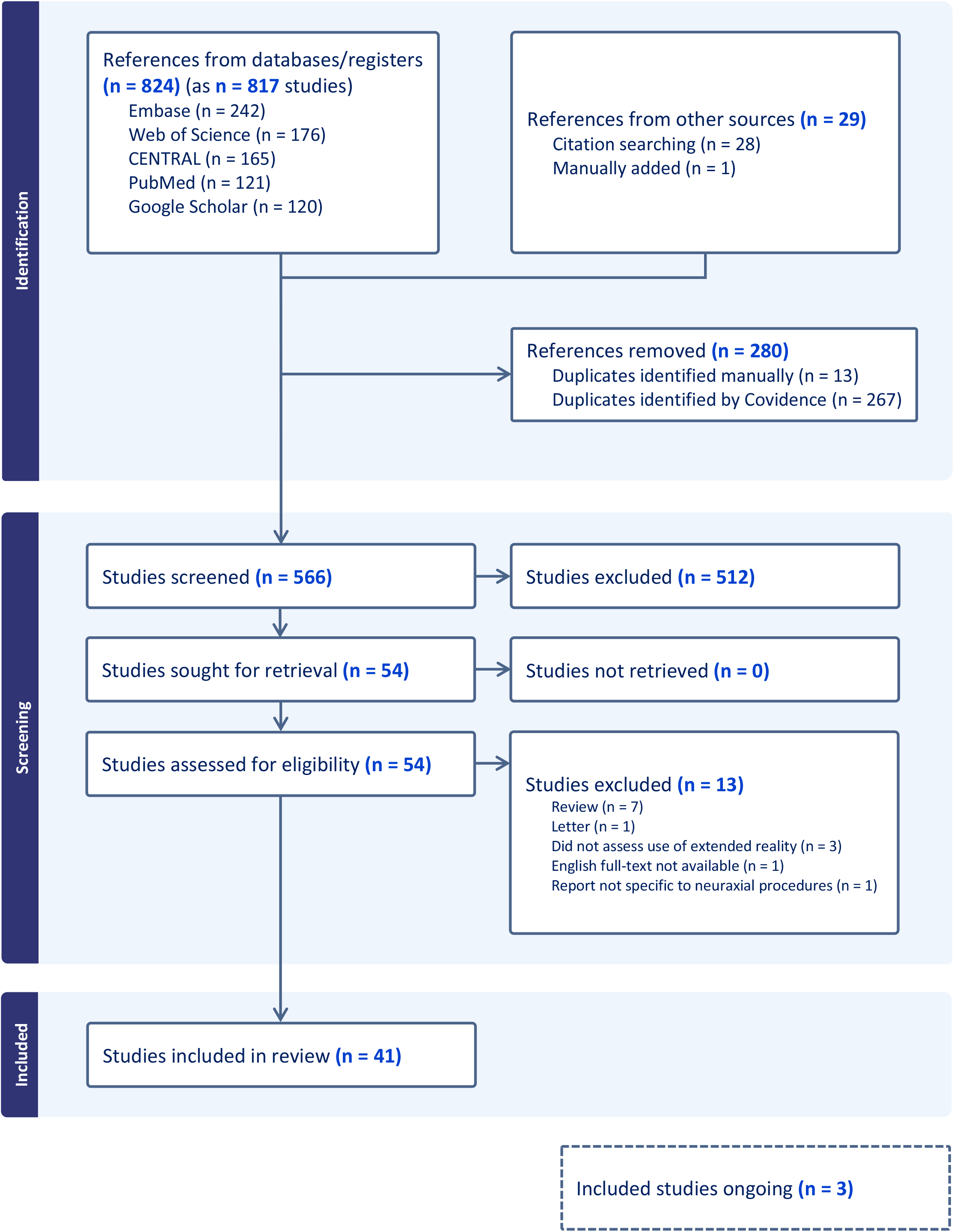
PRISMA flowchart of the review process.

A total of 13 articles were excluded.[4,9–20] Three were deemed irrelevant as XR was not utilized.[9–11] One article was not available in English and was excluded.[12] A letter describing the theoretical impact of XR on obstetric anesthesia was also excluded.[13] Seven reviews were excluded,[4,14–19] but relevant citations were incorporated into the screening pool. One article was excluded as the reported progress (i.e. viewing educational content using a head-mounted device) was not considered specific to teaching neuraxial procedures.[20]

The characteristics of the included evidence are displayed in **table 2**. Clinical use of XR was infrequent, with only four of 38 completed studies reporting the use of XR in patient care. Researchers from Canada and the United States were first authors for more than 50% of the included studies, but those from Europe, China, and South Korea have made substantial contributions. Both AR and VR were frequently used. Eighteen completed studies used computer screens to display XR visuals, and sixteen used head-mounted devices (HMDs). The remaining four studies employed either laser projectors or semi-transparent mirrors to project relevant visuals onto the subject’s back.

**Table 2.** Characteristics of included studies.

| Characteristic | Value | N (%) |
| --- | --- | --- |
| First author country | Canada | 11 (26.8) |
|  | United States | 10 (24.4) |
|  | China | 6 (14.6) |
|  | South Korea | 4 (9.8) |
|  | United Kingdom | 3 (7.3) |
|  | Germany | 2 (4.9) |
|  | Brazil | 1 (2.4) |
|  | Ireland | 1 (2.4) |
|  | Japan | 1 (2.4) |
|  | Portugal | 1 (2.4) |
|  | Singapore | 1 (2.4) |
| Year of publication | Before 2000 | 2 (4.9) |
|  | 2000-2010 | 1 (2.4) |
|  | 2011-2020 | 18 (43.9) |
|  | 2021-2024 | 20 (48.8) |
| Form of XR | AR/MR | 23 (56.1) |
|  | VR | 17 (41.5) |
|  | Both AR and VR | 1 (2.4) |
| Described use of XR<sup>a</sup> | Clinical use | 4 (10.5) |
|  | Preclinical development | 13 (34.2) |
|  | Educational | 21 (55.3) |
| Study design | RCTs | 5 (12.2) |
|  | Planned RCTs | 3 (7.3) |
|  | Non-RCT Experiments | 17 (41.5) |
|  | Case report/series | 4 (9.8) |
|  | Technical report / Usability | 12 (29.3) |
| XR device | Computer screen | 18 (43.9) |
|  | Head-Mounted Device | 16 (39) |
|  | Semi-transparent mirror | 2 (4.9) |
|  | Projector | 2 (4.9) |
|  | Unspecified | 3 (7.3) |
| Source of funding | Not reported | 9 (22) |
|  | Not sponsored | 4 (9.8) |
|  | Industry | 4 (9.8) |
|  | Public, department, or non-profit | 24 (58.5) |

Abbreviations: XR, extended reality; AR, augmented reality; MR, mixed reality; VR, virtual reality; RCT, randomized-controlled trial
a: excludes clinical trial pre-registrations

A summary of each study is provided in **supplemental table 1** and discussed below.

### Clinical applications of extended reality

Among the four studies detailing the clinical use of XR, our review identified only one randomized controlled trial (RCT) investigating XR navigation in patient care. Wiegelmann et al.[21] used a previously described technique[22] to overlay a holographic line representing the optimal needle trajectory from the skin to the thoracic epidural space. This holographic line was visible to proceduralists wearing an HMD as they performed a thoracic epidural. This study found a significant reduction in average procedure time with XR compared to traditional approaches (4.5 minutes vs. 7.3 minutes, p=0.02).[21] Although there was a significant reduction in the number of needle movements required (7.2 vs 14.4, p=0.01), there was no noticeable difference in the pain associated with the procedure.

VR has successfully been used for preprocedure simulation in challenging interventional procedures. A case report by Wang et al. describes the successful use of an existing VR anatomy application to determine the optimal fluoroscopic angulation in a patient with a suspected schwannoma undergoing a transforaminal epidural injection at the level of the lesion.[23] Seong et al. developed VR software enabling clinicians to simulate needle interventions using patient computed tomography (CT) scans.[24] This system allowed clinicians to simulate fluoroscopy and obtain x-ray images representative of the real patient to determine the ideal approach. Clinicians successfully executed a challenging transforaminal epidural injection following prior failed attempts by implementing the preprocedure plan designed in VR.[24]

AR was applied in remote consultations during spinal cord stimulator surgeries. Fritz et al. detail the use of AR goggles which enabled a remote specialist, located 200 miles away, to observe the procedure and provide annotated images and live commentary.[25] This led to the successful insertion of paddle leads in a patient with highly challenging anatomy.[25]

Our review includes three uncompleted clinical trial registrations, all filed by the same group of investigators and listed as “not yet recruiting” according to the Chinese Clinical Trial Registry.[26–28] The researchers aim to compare the success rates of first-pass lumbar punctures for clinicians using mixed reality guidance versus those using the conventional approach. Efforts to contact the researchers for updates on the studies were unsuccessful. A summary of the group’s previous work is provided later in this review.[29]

### Preclinical applications of extended reality

We identified thirteen preclinical investigations of XR primarily exploring how AR/MR technology could enhance or replace traditional navigation methods.

A significant focus was placed on improving ultrasound-assisted neuraxial procedures using AR and MR. Three studies utilized image processing algorithms to automatically detect and project lumbar spine levels[30,31] or the midline[32] during neuraxial ultrasound. A fourth study introduced a machine learning/artificial intelligence-based system for automatic detection and projection of spine levels.[33] The authors suggest such systems could be used clinically by anesthesiologists to determine the appropriate site of initial needle insertion.

Continuous ultrasound guidance during needle placement can present ergonomic and visual challenges. Ameri et al. reported a possible solution, using a tracked epidural needle and augmented reality (AR) to improve visualization of the needle and anatomy on ultrasound. This system showed a lower rate of accidental dural punctures compared to using only ultrasound in trials conducted on a phantom.[34] An alternative approach involved using a head-mounted display (HMD) to create a holographic line representing the optimal needle trajectory from the skin to the epidural space.[22] This method was validated in a randomized controlled trial (RCT), which is summarized earlier in this review.[21]

Fritz et al. addressed limitations of magnetic resonance imaging-guided lumbar spine interventions by overlaying preprocedural axial magnetic resonance images onto the procedural field using a semi-transparent mirror.[35,36] This system enabled successful epidural injections on cadavers without repeat scanning, with a median procedure time of 8.6 minutes.[36]

In a study with volunteers, Wu et al. assessed the accuracy of overlaying holograms of spinous processes on their actual locations.[29] Computed tomography (CT) images were used to generate 3-dimensional images of the spine, which were projected over the subject’s backs using an HMD.[29] Markers were placed over the holographic spinous processes, and the subjects underwent a repeat CT scan to assess the accuracy of marker placement. The maximum mean error along one axis was 4.2 mm.[29] A significant limitation of this system is that, for clinical use, patients would need to undergo preprocedural CT scanning while using a posture fixation device and maintain that position during the neuraxial procedure. The researchers plan to conduct a randomized controlled trial, and registrations were submitted to the Chinese Clinical Trial Registry as discussed previously in this review.[26–28]

In fluoroscopy-guided neuraxial procedures, AR showed promise in reducing radiation exposure without compromising accuracy. Reinacher et al. demonstrated high success rates with AR in simulated neuraxial access on prone phantoms[37]. On phantoms with simulated respiratory motion, AR combined with fluoroscopy reduced procedure time and radiation exposure.[38,39]

Finally, an atypical use case for XR neuraxial simulators was presented by Cometa et al.[40] They compared the epidural space overshoot associated with different loss-of-resistance techniques using measurements derived from an MR-based simulator. Intermittent needle advancement with intermittent plunger pressure resulted in the most significant overshoot.[40]

### Educational applications of extended reality

Early applications of XR for neuraxial education involved non-immersive VR, presenting a virtual patient on a computer screen for a simulated neuraxial procedure.[41–45] Haptic devices controlled the virtual needle and provided feedback as the needle traversed simulated tissues of varying density. Attempts to enhance realism incorporated force data derived from porcine models[41,42], CT data,[43] and MRI intensity levels[44]. In order to increase engagement, some iterations incorporated gamification, or the use of game design elements in non-gaming contexts,[46] by awarding points[43] and virtual “achievements”[47].

Kulcsár et al. conducted a pilot randomized-controlled trial using a non-immersive VR spinal anesthesia simulator, comparing it to low-fidelity training on an orange.[48] Although no significant differences were observed in multiple-choice and simulator-based tests, simulator-trained interns exhibited better performance on real patients according to a global rating scale.[48]

One advantage of XR is its ability to provide information that is typically unavailable. In one study, an interactive spine model was used to improve trainee knowledge through simulation of spinal ultrasound scanning.[49] Edwards et al. applied a mixed reality simulator to develop a modified paramedian thoracic epidural technique, which was taught to trainees and then applied in challenging patients.[50] In a 2015 RCT, Keri et al. found that residents who practiced ultrasound-guided lumbar puncture with AR enhancement outperformed those who practiced with ultrasound alone.[51]

Hologram-producing HMDs have found use in neuraxial education. Usability studies using HoloLens (Microsoft Corporations, Redmond, Washington) for training garnered positive responses from participants.[52–55] However, in an RCT conducted by Hayasaka et al., it failed to demonstrate a lasting difference between students trained on an AR-enhanced phantom and those trained without AR.[55]

Most recent VR applications were built using the Unity game engine (Unity Software Inc., San Francisco, California).[47,56–59] Unity simplifies the development of XR applications and reduces duplicate work by providing pre-built code.[60] In one RCT involving a Unity-based immersive VR simulator, trainees learning lumbar transforaminal epidural block placement using VR outperformed those who learned via text and video material alone.[58]

Although software like Unity has reduced the redundant work in the development of XR education and simulation software, there is significant heterogeneity in how individual systems are developed. Lampotang et al. argue that these heterogeneous approaches may lead to increased validation and curriculum development costs. They describe the System of Modular, Mixed and Augmented Reality Tracking Simulators (SMMARTS) as a potential solution.[61] SMMARTs enables the reuse of hardware and software components and allows for the development of varied procedural simulators, including an epidural loss-of-resistance simulator.[61]

Our search strategy identified one validation study of commercial XR simulators. White and Jung conducted a pre- and post-test study using a spinal cord stimulator training system.[62] After seven minutes of instruction using the VR simulator, participants had subjective and objective improvements in their performance.[62] A notable limitation was the $50,000 cost of the system (SPINE Mentor, Surgical Science, Göteborg, Sweden).[62]

## DISCUSSION

To our knowledge, this is the first scoping review on the uses of XR technology for neuraxial anesthesia and pain procedures. We identified 38 completed and three pending studies investigating the use of XR for neuraxial procedures. Our findings demonstrate accelerating interest in the use of XR technology. The first included article was published in 1996, but a significant majority was published after 2015, as shown in **figure 2**, as mainstream commercial XR devices were released. The release and widespread adoption of commercial, standalone devices such as the Meta Quest 3 (Meta Inc., Menlo Park, CA) and the Vision Pro (Apple Inc., Cupertino, CA) is expected to further drive XR research.

**Figure 2:**
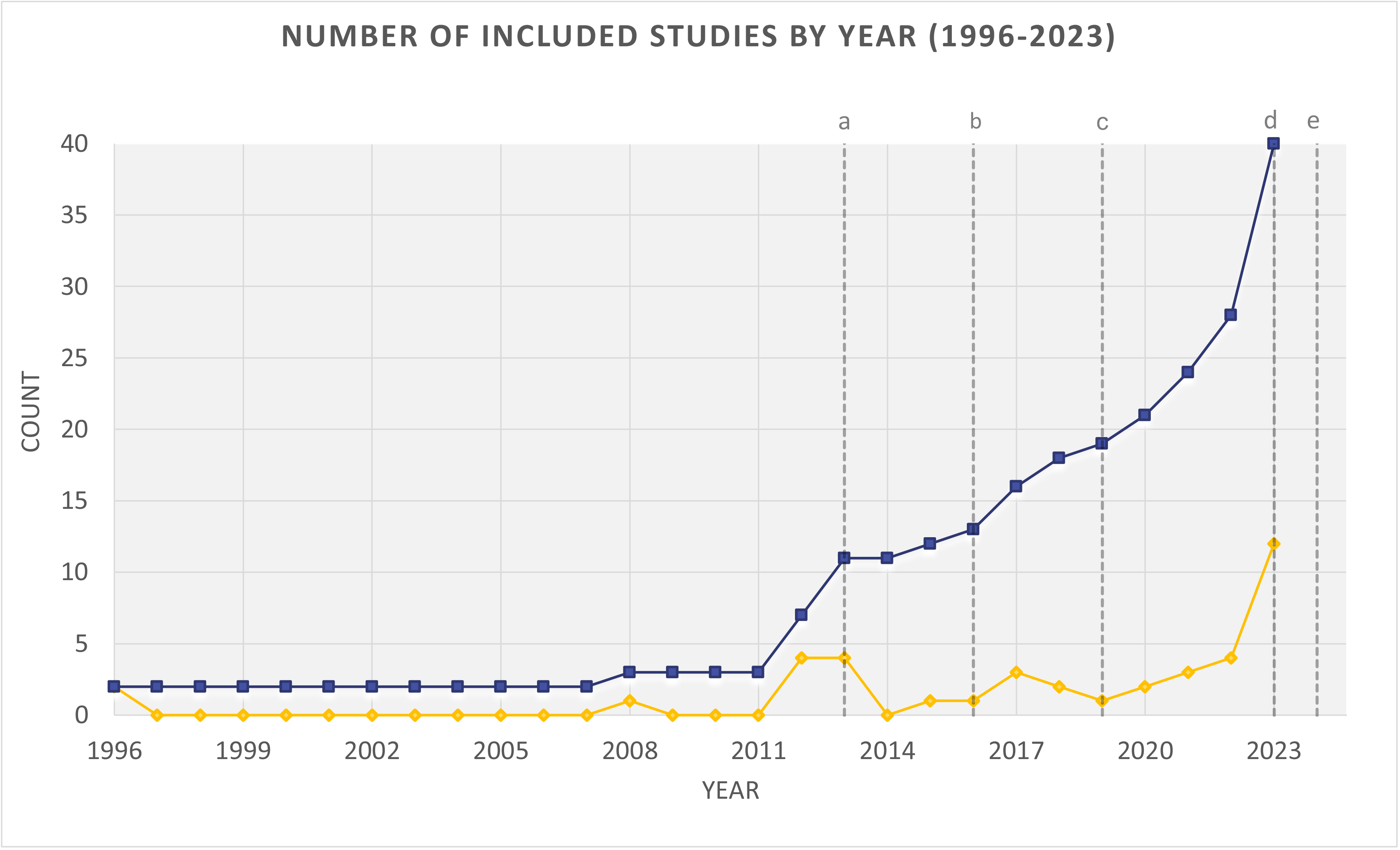
Number of included studies by year (1996 to 2023) a: Release of Oculus Rift DK1 b: Release of Microsoft HoloLens c: Release of Microsoft HoloLens 2 d: Release of Meta Quest 3 e: Release of Apple Vision Pro

While a significant number of studies have been conducted in Canada and the United States, our results indicate global interest in XR for neuraxial procedures. However, when accounting for duplicate authors, the total number of active research groups in this field remains limited.

To avoid confusion stemming from misuse of terminology, we intentionally grouped AR and MR in our analysis. AR, MR, and VR exist on a spectrum[60], and modern spatial computing devices often support multiple forms of XR. This makes functional categorization (e.g., simulation vs. procedural navigation) more relevant than the relative visibility of virtual and physical content.

A majority of the included studies evaluated the use of XR in educational settings. Surveyed trainees and clinicians displayed favorable responses to the adoption of XR for neuraxial education.[19,44,52,53,55,63] The largest reported survey of trainees and anesthesiologists included more than 90 respondents, and VR simulation was highly rated on various metrics, including ease of use (median score eight out of ten) and visual quality (median score 8 out of ten).[44] However, there is a lack of research into the clinical transferability of XR-based training.

Clinical use of XR for neuraxial procedures remains limited. This is in contrast to specialties such as neurosurgery, where AR is used clinically for intraoperative navigation.[64] Similar application of XR navigation would confer numerous benefits in neuraxial procedures. If patient-specific anatomic holograms can be precisely overlayed onto the procedural field, XR may allow intuitive and hands-free navigation (versus ultrasound), improve accuracy (versus blind procedures), and reduce radiation exposure (versus fluoroscopy).

Despite these potential benefits, certain barriers hinder the widespread adoption of XR technology for neuraxial navigation. The cost of XR systems can vary widely depending on system complexity, and how clinicians and patients will respond to non-traditional navigation modes is still unknown. However, the most significant barrier lies in displaying accurate navigational information for each individual patient. Preprocedural cross-sectional imaging is often unavailable for patients undergoing neuraxial procedures. Even when imaging is available, patient positioning can dramatically affect the shape of the spine, making image-to-patient registration challenging. Existing research suggests that it’s possible to incorporate XR into ultrasound-assisted neuraxial procedures with minimal impact on workflow[21]. However, clinical validation for fluoroscopic-guided procedures has not been performed.

Based on the results of this review, several unanswered questions emerge. Does XR guidance improve the safety and accuracy of neuraxial procedures? Are XR systems cost-effective for use in neuraxial procedures? Do the benefits of XR-enhanced simulators translate to better clinical performance? What role does machine learning play in registering preprocedural images onto real patients? How can the research community minimize the need for duplicate work in XR system development?

### Limitations

This scoping review has several limitations. While our search and screening criteria allowed for the inclusion of diverse studies published in various forms, there is likely to be publication bias in the included studies, many of which report favorable results. A formal assessment of the quality of the evidence is beyond the scope of a scoping review, and, as such, we do not provide practice recommendations.

Our study deviated slightly from the pre-registered protocol in a few instances. During the screening process, we decided to exclude studies not available in English, leading to the exclusion of one study. Additionally, we excluded reviews, although relevant citations were included. Originally, we did not anticipate the need to contact the authors of individual studies, but we ended up having to contact three authors to obtain full-text articles. We believe that these deviations had minimal or even positive impacts on the review.

Despite these limitations, this scoping review offers a comprehensive overview of the available research on the use of XR for neuraxial procedures. Our broad and iterative search strategy led to the screening of a large number of studies, and our inclusive eligibility criteria led to the inclusion of diverse studies spanning nearly three decades. As intended, this scoping review maps the existing evidence, identifies knowledge gaps, and provides suggestions for further research.

## CONCLUSIONS

This scoping review emphasizes the diverse applications of XR technology in the teaching and performance of neuraxial procedures within the fields of anesthesiology and pain medicine. Although many of the included studies were exploratory in nature, there is a growing cohort of researchers investigating the potential of XR in clinical settings. To advance the field, future research should prioritize investigations into the safety, accuracy, practicality, and cost-effectiveness of XR. A concerted effort in these areas is expected to catalyze adoption of this technology and enhance the quality of both education and patient care.

## Supporting information

PRISMA Checklist

## Data Availability

All data produced in the present work are contained in the manuscript

## REFERENCES

1. Co M, Chiu S, Billy Cheung HH. Extended reality in surgical education: A systematic review. Surgery. 2023 Nov;174(5):1175–83.

2. Zhang J, Lu V, Khanduja V. The impact of extended reality on surgery: a scoping review. Int Orthop. 2023 Mar;47(3):611.

3. Curran VR, Xu X, Aydin MY, Meruvia-Pastor O. Use of Extended Reality in Medical Education: An Integrative Review. Med Sci Educ. 2022 Dec 19;33(1):275–86.

4. Privorotskiy A, Garcia VA, Babbitt LE, Choi JE, Cata JP. Augmented reality in anesthesia, pain medicine and critical care: a narrative review. J Clin Monit Comput. 2022 Feb;36(1):33–9.

5. Kamimura Y, Yamamoto N, Shiroshita A, Miura T, Tsuji T, Someko H, et al. Comparative efficacy of ultrasound guidance or conventional anatomical landmarks for neuraxial puncture in adult patients: a systematic review and network meta-analysis. Br J Anaesth [Internet]. 2023 Oct 6 [cited 2023 Dec 14]; Available from: https://www.sciencedirect.com/science/article/pii/S0007091223005020

6. Manchikanti L, Knezevic NN, Navani A, Christo PJ, Limerick G, Calodney AK, et al. Epidural Interventions in the Management of Chronic Spinal Pain: American Society of Interventional Pain Physicians (ASIPP) Comprehensive Evidence-Based Guidelines. Pain Physician. 2021 Jan;24(S1):S27–208.

7. Cho J, Thaker D. Extended Reality for Neuraxial Anesthesia and Pain Procedures: A Scoping Review Protocol. 2023 Nov 5 [cited 2023 Dec 14]; Available from: https://osf.io/5u7zs

8. Tricco AC, Lillie E, Zarin W, O’Brien KK, Colquhoun H, Levac D, et al. PRISMA Extension for Scoping Reviews (PRISMA-ScR): Checklist and Explanation. Ann Intern Med. 2018 Oct 2;169(7):467–73.

9. Kerby B, Rohling R, Nair V, Abolmaesumi P. Automatic identification of lumbar level with ultrasound. Annu Int Conf IEEE Eng Med Biol Soc IEEE Eng Med Biol Soc Annu Int Conf. 2008;2008:2980–3.

10. Lee RA, van Zundert TCRV, van Koesveld JJM, van Zundert A a. J, Stolker RJ, Wieringa PA, et al. Evaluation of the Mediseus epidural simulator. Anaesth Intensive Care. 2012 Mar;40(2):311–8.

11. Novak-Jankovič V. Simulation-Based Training of Regional Anesthesia. Acta Clin Croat. 2022 Sep 1;61.(Supplement 2):155–9.

12. Gomes DC, Machado LS. A Simulator for Regional Anesthesia Training. In: 2017 19th Symposium on Virtual and Augmented Reality (SVR) [Internet]. 2017 [cited 2024 Jan 14]. p. 289–92. Available from: https://ieeexplore.ieee.org/document/8114450

13. Rubin JE, White RS, Boyer RB, Jotwani R. Obstetric anesthesiology and extended reality: an introduction to future uses of spatial computing in obstetric anesthesiology. Int J Obstet Anesth [Internet]. 2023 [cited 2023 Dec 14]; Available from: https://www.obstetanesthesia.com/article/S0959-289X(23)00313-8/abstract

14. Kumar AH, Sultan E, Mariano ER, Udani AD. A modern roadmap for the use of simulation in regional anesthesiology training. Curr Opin Anaesthesiol. 2022;35(5):654–9.

15. Vaughan N, Dubey VN, Wee MY, Isaacs R. A review of epidural simulators: where are we today? Med Eng Phys. 2013;35(9):1235–50.

16. Sauer F, Vogt S, Khamene A. Augmented Reality. In: Peters T, Cleary K, editors. Image-Guided Interventions: Technology and Applications [Internet]. Boston, MA: Springer US; 2008 [cited 2024 Jan 13]. p. 81–119. Available from: 10.1007/978-0-387-73858-1_4

17. Dubey V, Vaughan N, Wee MY, Isaacs R, Andrade A. Biomedical engineering in epidural anaesthesia research [Internet]. InTech London, UK; 2013 [cited 2023 Dec 14]. Available from: 10.5772/50764

18. Elsharkawy H, Sonny A, Chin KJ. Localization of epidural space: A review of available technologies. J Anaesthesiol Clin Pharmacol. 2017;33(1):16.

19. Ramlogan RR, Chuan A, Mariano ER. Contemporary training methods in regional anaesthesia: fundamentals and innovations. Anaesthesia. 2021 Jan;76(S1):53–64.

20. Sargsyan N, Bassy R, Wei X, Akula S, Liu J, Seals C, et al. Augmented Reality Application to Enhance Training of Lumbar Puncture Procedure: Preliminary Results. In: EPiC Series in Computing [Internet]. EasyChair; 2019 [cited 2024 Jan 13]. p. 189–96. Available from: https://easychair.org/publications/paper/3dm5

21. Wiegelmann J, Choi S, McHardy PG, Matava C, Singer O, Kaustov L, et al. Randomized control trial of a holographic needle guidance technique for thoracic epidural placement. Reg Anesth Pain Med. 2024 Jan 11;rapm-2023-104703.

22. Tanwani J, Alam F, Matava C, Choi S, McHardy P, Singer O, et al. Development of a Head-Mounted Holographic Needle Guidance System for Enhanced Ultrasound-Guided Neuraxial Anesthesia: System Development and Observational Evaluation. JMIR Form Res. 2022;6(6):e36931.

23. Wang EF, Yu J, Rubin JE, Jotwani R. Virtual reality training and modeling to aid in pre-procedural practice for thoracic nerve root block in the setting of a schwannoma. Interv Pain Med. 2023 Mar 1;2(1):100180.

24. Seong H, Yun D, Yoon KS, Kwak JS, Koh JC. Development of pre-procedure virtual simulation for challenging interventional procedures: an experimental study with clinical application. Korean J Pain. 2022 Oct 1;35(4):403–12.

25. Fritz AK, Steele N, Patel C. Augmented Reality Technology in Neuromodulation Surgery. Neuromodulation. 2022 Oct 1;25(7):S255–6.

26. Gu, W. Development and application of mixed reality guided spinal puncture. 2017; Available from: https://www.chictr.org.cn/showprojEN.html?proj=18023

27. Lei, G. Mixed reality-assisted Versus Landmark-guided spinal puncture in elderly patients: a randomized controlled pilot study. 2023; Available from: https://www.chictr.org.cn/showprojEN.html?proj=178960

28. Lei, G, Gu, W. Mixed reality-assisted Versus Landmark-guided spinal puncture in elderly patients: a stratified randomized controlled trial. 2023; Available from: https://www.chictr.org.cn/showprojEN.html?proj=189622

29. Wu J, Gao L, Shi Q, Qin C, Xu K, Jiang Z, et al. Accuracy Evaluation Trial of Mixed Reality-Guided Spinal Puncture Technology. Ther Clin Risk Manag. 2023;19:599–609.

30. Ashab HAD, Lessoway VA, Khallaghi S, Cheng A, Rohling R, Abolmaesumi P. An augmented reality system for epidural anesthesia (AREA): prepuncture identification of vertebrae. IEEE Trans Biomed Eng. 2013;60(9):2636–44.

31. Ashab HAD, Lessoway VA, Khallaghi S, Cheng A, Rohling R, Abolmaesumi P. AREA: an augmented reality system for epidural anaesthesia. Annu Int Conf IEEE Eng Med Biol Soc. 2012;2012:2659–63.

32. Toews A, Massey S, Gunka V, Lessoway V, Rohling R. ProjectAlign: a real-time ultrasound guidance system for spinal midline detection during epidural needle placement. In 2018.

33. Hetherington J, Lessoway V, Gunka V, Abolmaesumi P, Rohling R. SLIDE: automatic spine level identification system using a deep convolutional neural network. Int J Comput Assist Radiol Surg. 2017;12(7):1189–98.

34. Ameri G, Rankin A, Baxter JSH, Moore J, Ganapathy S, Peters TM, et al. Development and Evaluation of an Augmented Reality Ultrasound Guidance System for Spinal Anesthesia: Preliminary Results. Ultrasound Med Biol. 2019;45(10):2736–46.

35. Fritz J, U-Thainual P, Ungi T, Flammang A, Cho N, Fichtinger G, et al. Augmented Reality Visualization With Image Overlay for MRI-Guided Intervention: Accuracy for Lumbar Spinal Procedures With a 1.5-T MRI System. Am J Roentgenol. 2012;198(3):W266–73.

36. Fritz J, U-Thainual P, Ungi T, Flammang A, Fichtinger G, Iordachita I, et al. Augmented reality visualisation using an image overlay system for MR-guided interventions: technical performance of spine injection procedures in human cadavers at 1.5 Tesla. Eur Radiol. 2013;23(1):235–45.

37. Reinacher PC, Cimniak A, Demerath T, Schallner N. Usage of augmented reality for interventional neuraxial procedures: A phantom-based study. Eur J Anaesthesiol. 2023;40(2):121–9.

38. Lim S, Ha J, Yoon S, Sohn Y, Seo J, Koh J, et al. Augmented Reality Assisted Surgical Navigation System for Epidural Needle Intervention. In 2021. p. 4705–8.

39. Jun EK, Lim S, Seo J, Lee KH, Lee JH, Lee D, et al. Augmented Reality-Assisted Navigation System for Transforaminal Epidural Injection. J Pain Res. 2023;16:921–31.

40. Cometa MA, Lopez BM, Vasilopoulos T, Destephens AJ, Bigos A, Lizdas DE, et al. Does the Technique for Assessing Loss of Resistance Alter the Magnitude of Epidural Needle Tip Overshoot? Simul Healthc J Soc Simul Healthc. 2020 Jun;15(3):154–9.

41. Stredney D, Sessanna D, McDonald J, Hiemenz L, Rosenberg L. A virtual simulation environment for learning epidural anesthesia. In 1996. p. 164–75.

42. Vaughan N, Dubey VN, Wee MYK, Isaacs R. Advanced Epidural Simulator with 3D Flexible Spine and Haptic Interface. J Med Devices [Internet]. 2012;6(017524). Available from: 10.1115/1.4026702

43. Färber M, Hoeborn E, Dalek D, Hummel F, Gerloff C, Bohn CA, et al. Training and evaluation of lumbar punctures in a VR-environment using a 6DOF haptic device. Stud Health Technol Inform. 2008;132:112–4.

44. Hiemenz L, McDonald J, Stredney D, Sessanna D. A physiologically valid simulator for training residents to perform an epidural block. In 1996. p. 170–3. Available from: https://ieeexplore.ieee.org/abstract/document/493141/citations#citations

45. Vaughan N, Dubey VN, Wee MYK, Isaacs R. Virtual Reality Based Enhanced Visualization of Epidural Insertion. In American Society of Mechanical Engineers Digital Collection; 2013. p. 1413–8. Available from: 10.1115/DETC2012-70951

46. Deterding S, Khaled R, Nacke L, Dixon D. Gamification: Toward a definition. CHI 2011 Gamification Workshop Proceedings. In: 2011 Annual Conference on Human Factors in Computing Systems (CHI’11). 2011.

47. Brazil AL, Conci A, Clua E, Bittencourt LK, Baruque LB, Silva Conci N da. Haptic forces and gamification on epidural anesthesia skill gain. Entertain Comput. 2018 Mar 1;25:1–13.

48. Kulcsar Z, Aboulafia A, Hall T, Shorten GD. Determinants of learning to perform spinal anaesthesia: a pilot study. Eur J Anaesthesiol. 2008 Dec;25(12):1026–31.

49. Ramlogan R, Niazi AU, Jin R, Johnson J, Chan VW, Perlas A. A Virtual Reality Simulation Model of Spinal Ultrasound: Role in Teaching Spinal Sonoanatomy. Reg Anesth Pain Med. 2017;42(2):217–22.

50. Edwards DA, Vazquez R, Lizdas D, Lampotang S. A mixed reality simulator augmented with real-time 3D visualization helps develop a modified technique for accessing the thoracic epidural space. Reg Anesth Pain Med [Internet]. 2016;41(5). Available from: https://www.embase.com/search/results?subaction=viewrecord&id=L614572639&from=export

51. Keri Z, Sydor D, Ungi T, Holden MS, McGraw R, Mousavi P, et al. Computerized training system for ultrasound-guided lumbar puncture on abnormal spine models: a randomized controlled trial. Can J Anesth. 2015;62(7):777–84.

52. da Silva D, Costa CB, da Silva NA, Ventura I, Leite FP, Lopes DS. Augmenting the training space of an epidural needle insertion simulator with HoloLens. Comput Methods Biomech Biomed Eng Imaging Vis. 2022;10(3):260–5.

53. Lau WK, Chan JJI, Chan CLJ, Tan CW, Sng BL. Use of Augmented Reality in Learning Lumbar Spinal Anatomy for Training in Labor Epidural Insertion: A Pilot Study. Bali J Anesthesiol. 2023;7(3):135–40.

54. Huang X, Yan Z, Gong C, Zhou Z, Xu H, Qin C, et al. A mixed-reality stimulator for lumbar puncture training: a pilot study. BMC Med Educ. 2023 Mar 22;23(1):178.

55. Hayasaka T, Kawano K, Onodera Y, Suzuki H, Nakane M, Kanoto M, et al. Comparison of accuracy between augmented reality/mixed reality techniques and conventional techniques for epidural anesthesia using a practice phantom model kit. BMC Anesth. 2023;23(1):171.

56. Shewaga R, Uribe-Quevedo A, Kapralos B, Alam F. A Comparison of Seated and Room-Scale Virtual Reality in a Serious Game for Epidural Preparation. IEEE Trans Emerg Top Comput. 2020;8(1):218–32.

57. Moo-Young J, Weber TM, Kapralos B, Quevedo A, Alam F. Development of Unity Simulator for Epidural Insertion Training for Replacing Current Lumbar Puncture Simulators. Cureus. 2021;13(2):e13409.

58. Kim JY, Lee JS, Lee JH, Park YS, Cho J, Koh JC. Virtual reality simulator’s effectiveness on the spine procedure education for trainee: a randomized controlled trial. Korean J Anesth. 2023;76(3):213–26.

59. Zheng T, Xie H, Gao F, Gong C, Lin W, Ye P, et al. Research and application of a teaching platform for combined spinal-epidural anesthesia based on virtual reality and haptic feedback technology. BMC Med Educ. 2023;23(1):794.

60. Unity - Manual: XR [Internet]. Unity Documentation. [cited 2024 Jan 19]. Available from: https://docs.unity3d.com/Manual/XR.html

61. Lampotang S, Bigos AK, Avari K, Johnson WT, Mei V, Lizdas DE. SMMARTS: An Open Architecture Development Platform for Modular, Mixed, and Augmented Reality Procedural and Interventional Simulators. Simul Heal. 2021;16(5):353–61.

62. White WW Jr, Jung MJ. Three-Dimensional Virtual Reality Spinal Cord Stimulator Training Improves Trainee Procedural Confidence and Performance. Neuromodulation. 2023;26(7):1381–6.

63. Kagalwala DZ, Adhikary S, Murray WB, Webster R. The use of a computerized haptic simulation model to track angles of epidural needle insertion by anesthesiology residents. Br J Anaesth. 2012;108((Kagalwala D.Z.; Adhikary S.; Murray W.B.; Webster R.) Pennsylvania State University, College of Medicine, Hershey, PA, United States):ii179–80.

64. Cannizzaro D, Zaed I, Safa A, Jelmoni AJM, Composto A, Bisoglio A, et al. Augmented Reality in Neurosurgery, State of Art and Future Projections. A Systematic Review. Front Surg. 2022 Mar 11;9:864792.

