## Supplemental Table 1 for "Extended Reality for Neuraxial Anesthesia and Pain Procedures: A Scoping Review"

**Supplemental Table 1 Legend**

| 2D - Two-Dimensional  3D - Three-Dimensional  AR - Augmented Reality  CAAs - Certified Anesthesiologist Assistants  CSE - Combined Spinal-Epidural  CRNAs - Certified Registered Nurse Anesthetists  CT - Computed Tomography  DOF - Degree-of-freedom  LOR - Loss of Resistance  MRI - Magnetic Resonance Imaging  MR - Mixed Reality  N/A - Not Applicable  RCT - Randomized Control Trial | RMS - Root Mean Square  SMMARTS - Simulation and Modeling in Mixed and Augmented Reality Training Systems  US - Ultrasound  VR - Virtual Reality  XR - Extended Reality |
| --- | --- |

**Supplemental Table 1:** Results of individual sources of evidence

| **Authors (Year)** | **Title** | **Study design** | **Mode of XR** | **XR Display Device** | **Procedure** | **Participants** |
| --- | --- | --- | --- | --- | --- | --- |
| ***Clinical applications of extended reality*** | | | | | | |
| Gu (2017) | Development and application of mixed reality guided spinal puncture | Planned RCT | AR/MR | Unspecified | Lumbar puncture | 100 patients |
| Fritz et al. (2022) | Augmented reality technology in neuromodulation surgery | Case report | AR/MR | AR Goggles (Vuzix, Rochester, New York) | Revision spinal cord stimulator surgery with remote specialist consultation | 1 patient undergoing revision spinal cord stimulator surgery |
| Seong et al. (2022) | Development of pre-procedure virtual simulation for challenging interventional procedures: an experiments study with clinical application | Case report | VR | Unspecified | Fluoroscopy-guided transforaminal epidural injection | One 84-year-old woman with degenerative spine disease and several prior failed attempts at intervention |
| Lei (2023) | Mixed reality-assisted Versus Landmark-guided spinal puncture in elderly patients: a randomized controlled pilot study | Planned RCT | AR/MR | HoloLens (Microsoft Corporation, Redmond, Washington) | Lumbar puncture | 18 patients |
| Lei and Gu (2023) | Mixed reality-assisted Versus Landmark-guided spinal puncture in elderly patients: a stratified randomized controlled trial | Planned RCT | AR/MR | Unspecified | Lumbar puncture | 84 patients |
| Wang et al. (2023) | Virtual reality training and modeling to aid in pre-procedural practice for thoracic nerve root block in the setting of a schwannoma | Case report | VR | Oculus Quest 2 (Meta Inc., Menlo Park, CA) | Fluoroscopy-guided transforaminal epidural injection | One 64-year-old woman with suspected schwannoma in the right T11/T12 foramen |
| Wiegelmann et al. (2024) | Randomized control trial of a holographic needle guidance technique for thoracic epidural placement | RCT | AR/MR | HoloLens | Thoracic epidural | 83 patients undergoing thoracic epidural placement |
| ***Educational applications of extended reality*** | | | | | | |
| Hiemenz et al. (1996) | A physiologically valid simulator for training residents to perform an epidural block | Usability study | VR | Computer screen | Lumbar epidural | >90 anesthesiologists and anesthesiology residents |
| Stredney et al. (1996) | A virtual simulation environment for learning epidural anesthesia | Technical report | VR | Computer screen and shutter glasses | Lumbar epidural | 3 anesthesia residents |
| Färber et al. (2008) | Training and evaluation of lumbar punctures in a VR-environment using a 6DOF haptic device | Usability study | VR | Computer screen with shutter glasses | Simulated lumbar puncture | "Several" with varying medical experience |
| Kagalwala et al. (2012) | The use of a computerized haptic simulation model to track angles of epidural needle insertion by anesthesiology residents | Non-RCT experiment | VR | Computer screen | Simulated epidural needle navigation on a virtual coin | 16 anesthesia residents |
| Vaughan et al. (2012) | Advanced Epidural Simulator with 3D Flexible Spine and Haptic Interface | Usability study | VR | Computer monitor | Simulated thoracic and lumbar epidural | Unspecified |
| Kulcsár et al. (2013) | Preliminary evaluation of a virtual reality-based simulator for learning spinal anesthesia. | RCT | VR | Computer screen | Spinal anesthesia | 27 medical interns with minimal spinal anesthesia/dural puncture experience |
| Vaughan et al. (2013) | Virtual Reality Based Enhanced Visualization of Epidural Insertion | Usability study | VR | Computer screen with Z800 3D visor (eMagin Corporation, Hopewell Junction, New York) | Epidural placement | Unspecified number of experienced anesthetists |
| Keri et al. (2015) | Computerized training system for ultrasound-guided lumbar puncture on abnormal spine models: a randomized controlled trial | RCT | AR/MR | Computer screen | Ultrasound-guided lumbar puncture on phantom | 24 residents from anesthesia and surgical programs |
| Edwards et al. (2016) | A mixed reality simulator augmented with real-time 3D visualization helps develop a modified technique for accessing the thoracic epidural space | Case series | AR/MR | Computer screen | Modified paramedian thoracic epidural technique | 10 anesthesia residents, 15 patients |
| Ramlogan et al. (2017) | A Virtual Reality Simulation Model of Spinal Ultrasound: Role in Teaching Spinal Sonoanatomy. | Non-RCT experiment | VR | Computer screen | Ultrasound-guided lumbar neuraxial procedures | 14 trainees with no prior experience with neuraxial ultrasound |
| Brazil et al. (2018) | Haptic forces and gamification on epidural anesthesia skill gain | Technical report | VR | Computer screen | Simulated lumbar epidural | 1 experienced physician |
| Shewaga et al. (2020) | A Comparison of Seated and Room-Scale Virtual Reality in a Serious Game for Epidural Preparation | Non-RCT experiment | VR | HTC Vive (HTC Corporation, New Taipei, Taiwan) | Simulated Epidural preparation (pre-placement steps) | 38 university students and 2 university staff |
| Lampotang et al. (2021) | SMMARTS: An Open Architecture Development Platform for Modular, Mixed, and Augmented Reality Procedural and Interventional Simulators. | Technical report | AR/MR | Computer screen | Simulated epidural (and others) | N/A |
| Moo-Young et al. (2021) | Development of Unity Simulator for Epidural Insertion Training for Replacing Current Lumbar Puncture Simulators | Technical report | VR | Computer screen | Lumbar epidural | N/A |
| da Silva et al. (2022) | Augmenting the training space of an epidural needle insertion simulator with HoloLens | Usability study | AR/MR | HoloLens | Lumbar epidural on mannequin (midline or paramedian) | 5 anesthesiology specialists, 1 intern |
| Hayasaka et al. (2023) | Comparison of accuracy between augmented reality/mixed reality techniques and conventional techniques for epidural anesthesia using a practice phantom model kit. | RCT | AR/MR | HoloLens | Paramedian lumbar epidural on phantom | 30 medical students with no previous experience performing epidural anesthesia |
| Huang et al. (2023) | A mixed-reality stimulator for lumbar puncture training: a pilot study. | Usability study | AR/MR | HoloLens | Lumbar puncture | 40 participants (students, faculty, interns, residents, nurse anesthetists, fellows) |
| Kim et al. (2023) | Virtual reality simulator's effectiveness on the spine procedure education for trainee: a randomized controlled trial. | RCT | VR | Oculus Quest 2 | Fluoroscopy-guided lumbar transforaminal epidural block | 20 first- or second-year residents inexperienced in C-arm use and undergoing training in anesthesiology and pain medicine |
| Lau et al. (2023) | Use of Augmented Reality in Learning Lumbar Spinal Anatomy for Training in Labor Epidural Insertion: A Pilot Study | Usability study | AR/MR | HoloLens | Intended to be used for lumbar neuraxial procedures, but only visualized anatomy in 3D as overlays on mannequins | 31 anesthesia specialists and nonspecialists (residents, medical officers) [Singapore system] |
| White and Jung (2023) | Three-Dimensional Virtual Reality Spinal Cord Stimulator Training Improves Trainee Procedural Confidence and Performance. | Non-RCT experiment | VR | Computer screen | Simulated spinal cord stimulator training | 14 pain, physical medicine, and anesthesiology trainees |
| Zheng et al. (2023) | Research and application of a teaching platform for combined spinal-epidural anesthesia based on virtual reality and haptic feedback technology. | Non-RCT experiment | VR | HTC Vive | Combined spinal epidural | 20 anesthesiology interns |
| ***Preclinical applications of extended reality*** | | | | | | |
| Ashab et al. (2012) | AREA: an augmented reality system for epidural anaesthesia. | Non-RCT experiment | AR/MR | Computer screen | Automated detection of lumbar spine levels during US-assisted lumbar neuraxial procedure | 10 volunteers |
| Fritz et al. (2012) | Augmented Reality Visualization With Image Overlay for MRI-Guided Intervention: Accuracy for Lumbar Spinal Procedures With a 1.5-T MRI System | Non-RCT experiment | AR/MR | Semi-transparent mirror | MRI-guided lumbar spine procedures on a human lumbar spine phantom (cadaveric donor) | N/A |
| Ashab et al. (2013) | An augmented reality system for epidural anesthesia (AREA): prepuncture identification of vertebrae. | Non-RCT experiment | AR/MR | Computer screen | Automated detection of lumbar spine levels during US-assisted lumbar neuraxial procedure | 17 volunteers |
| Fritz et al. (2013) | Augmented reality visualisation using an image overlay system for MR-guided interventions: technical performance of spine injection procedures in human cadavers at 1.5 Tesla | Non-RCT experiment | AR/MR | Semi-transparent mirror | MRI-guided lumbar spine injection procedures | 12 non-embalmed, full-torso human cadavers |
| Hetherington et al. (2017) | SLIDE: automatic spine level identification system using a deep convolutional neural network. | Non-RCT experiment | AR/MR | PicoPro laser projector (Celluon Inc., Seoul, South Korea) | Ultrasound-assisted lumbar neuraxial procedure | 20 participants |
| Toews et al. (2018) | ProjectAlign: a real-time ultrasound guidance system for spinal midline detection during epidural needle placement | Non-RCT experiment | AR/MR | PicoPro | Automated detection of midline during ultrasound-assisted neuraxial procedure | 12 volunteers |
| Ameri et al. (2019) | Development and Evaluation of an Augmented Reality Ultrasound Guidance System for Spinal Anesthesia: Preliminary Results. | Non-RCT experiment | AR/MR | Computer screen | US-guided lumbar epidural on phantom | 4 novice users and 1 expert anesthesiologist |
| Cometa et al. (2020) | Does the Technique for Assessing Loss of Resistance Alter the Magnitude of Epidural Needle Tip Overshoot? | Non-RCT experiment | AR/MR | Computer screen | Simulated epidural placement with 3 different LOR techniques | 45 anesthesia clinicians (trainees, CAAs, CRNAs, and anesthesiologists) |
| Lim et al. (2021) | Augmented Reality Assisted Surgical Navigation System for Epidural Needle Intervention | Non-RCT experiment | AR/MR | HoloLens | Fluoroscopy-guided epidural needle insertion on phantom | 1 pain physician |
| Tanwani et al. (2022) | Development of a Head-Mounted Holographic Needle Guidance System for Enhanced Ultrasound-Guided Neuraxial Anesthesia: System Development and Observational Evaluation. | Usability study | AR/MR | HoloLens | Ultrasound-assisted lumbar neuraxial anesthesia | 7 anesthesia residents and attendings |
| Jun et al. (2023) | Augmented Reality-Assisted Navigation System for Transforaminal Epidural Injection. | Non-RCT experiment | VR and AR/MR | HoloLens | Fluoroscopy-guided lumbosacral transforaminal epidural injection on torso phantom with simulated respirations | 1 anesthesiologist with 5-year experience in pain medicine |
| Reinacher et al. (2023) | Usage of augmented reality for interventional neuraxial procedures: A phantom-based study. | Non-RCT experiment | AR/MR | Magic Leap 1 (Magic Leap, Inc., Plantation, Florida) | Paramedian lower thoracic / upper lumbar epidural access on a phantom | 4 total: two anesthesiologists, one neuroradiologist, and one stereotactic neurosurgeon |
| Wu et al. (2023) | Accuracy Evaluation Trial of Mixed Reality-Guided Spinal Puncture Technology. | Non-RCT experiment | AR/MR | HoloLens | Lumbar puncture | 12 volunteers (average age 19.5) |

**Supplemental Table 1 (continued)**

| **Authors (Year)** | **Key findings** | **Participant response to XR use** | **Limitation of XR system** | **Suggestions for further research by authors** |
| --- | --- | --- | --- | --- |
| ***Clinical applications of extended reality*** | | | | |
| Gu (2017) | Study pending | Study pending | Study pending | Study pending |
| Fritz et al. (2022) | The technology enabled remote specialist support resulting in successful procedure in patient with challenging anatomy | Not reported | Non-described | None stated |
| Seong et al. (2022) | Procedure, including fluoroscopic navigation, simulated using VR. Technique replicated during actual procedure, which was successful. | Not reported | CTs performed supine, whereas procedure generally performed prone. This may affect accuracy. Virtual x-ray image quality can decrease at certain angles. | Application of similar simulators in other fields |
| Lei (2023) | Study pending | Study pending | Study pending | Study pending |
| Lei and Gu (2023) | Study pending | Study pending | Study pending | Study pending |
| Wang et al. (2023) | Preprocedure planning performed using VR to determine optimal fluoroscopic angulation. Angulation recreated during successful procedure. | Not reported | Described system does not allow importing of patient-specific images, and procedural models must be manually created | Additional use of VR for interventional planning for individualized cases |
| Wiegelmann et al. (2024) | Hologram group required half the time and fewer needle movements to complete a thoracic epidural, compared to those using conventional methods. No differences in procedure-related pain were noted. | Not reported | Did not explicitly get compared to ultrasound without MR (only 1 instance of preprocedural US use in control group) | Assess whether improved procedural efficiency improves patient outcomes |
| ***Educational applications of extended reality*** | | | | |
| Hiemenz et al. (1996) | Surveyed groups rated the VR simulator highly on various metrics (e.g. ease of use and visuals), with an average score of 7.17 out of 10 for all questions on survey. | See previous column | Device used for system development no longer commercially available | Improve haptic feedback to more realistically model human back, add saline filled plunger, incorporate paramedian approach, and improve resolution of MRI images used to create virtual spine |
| Stredney et al. (1996) | Porcine specimen used to measure force data during epidural placement to develop an epidural simulator with realistic haptic feedback. Qualitative study was performed to collect feedback from residents on system. | No specific comments on use of VR | Device used for system development no longer commercially available | Incorporate ability to test for LOR |
| Färber et al. (2008) | Users able to identify different tissue layers based on tactile feedback. Evaluation system gave feedback, and trainees were motivated to receive the "high score." | Not reported | None described | Incorporation of other data sets |
| Kagalwala et al. (2012) | Assessed whether residents would use smaller angle for epidural needle redirections after training with VR system. No changes noted post-training. | System may be useful for training | A part-task trainer without real clinical correlate | Further development of training device |
| Vaughan et al. (2012) | Subjectively realistic and accurate forces generated, allows practice on various simulated patients | Not reported | None specified | None specified |
| Kulcsár et al. (2013) | No significant differences on written test scores or on simulator-based testing for those trained on VR vs. low-fidelity simulator. Subset of two groups were evaluated on performance of spinal anesthesia in real patients. Those from the simulation group received higher scores on the global rating scale. | XR simulation adds realism at low stress levels | Bulky device compared to modern head-mounted devices | Record motion of participant for assessment; replicate complex scenarios with various levels of difficulty and variations |
| Vaughan et al. (2013) | Describes development of 3D graphics system for visualization of simulated epidural procedures | Subjects reported positive opinions about the use of stereo glasses for visualizing medical simulation. | None specified | None stated |
| Keri et al. (2015) | Group trained using AR software performed better than conventional US group on a post-test in terms of needle path, potential tissue damage, and time to needle insertion. Total procedure time and success rates were not statistically different | Not reported | Requires tracking hardware; phantom model not realistic | Evaluate whether skills developed using this system transfer to clinical domain |
| Edwards et al. (2016) | MR simulation allowed authors to develop a new approach to paramedian epidural placement allowing for better tactile feedback. This technique was "taught by the authors to 10 residents who successfully translated the technique and obtained the thoracic epidural space in 15 challenging patients.' | Not reported | Not described | Further evaluate the modified paramedian approach and the role of MR to enhance learning |
| Ramlogan et al. (2017) | Describes online interactive educational model that allows simulated ultrasound. 1 hour of self-study on this model led to almost a 40% improvement in scores on a previously validated test that can distinguish novices and experts. | Easy to use and beneficial to training | Does not provide haptic feedback | Assess clinical transferability and add varied spine models |
| Brazil et al. (2018) | Describes the development of a VR simulator with haptic feedback and a gamification strategy involving points and achievements to motivate and guide trainees | VR simulation might be "important and useful for medical skill" | Lack of immersion. LOR is not simulated on the haptic device. | Validate the impacts of gamification and haptic feedback |
| Shewaga et al. (2020) | Participants played a serious game involving preparation for an epidural placement in both seated and room-scale VR conditions. Participants' experiences were analyzed. | Participants responded positively, particularly with room-scale VR | Does not simulate placement of epidural needle | Include non-player characters to interact with and incorporate technical aspects of epidural placement. |
| Lampotang et al. (2021) | Describes the design of an open architecture development platform for 9 procedural simulators, including an epidural simulator. | N/A | System cost | Share solutions that reduce duplication of work |
| Moo-Young et al. (2021) | Describes development of VR epidural simulator using commercial-grade technologies. System has ability to simulate LOR using syringe. | N/A | Occasional graphical glitches reported. Does not provide a way to palpate a simulated patient's back. Use of a 2D screen reduces immersion. | Validate system, improve immersion, and add gamification features. |
| da Silva et al. (2022) | Prototype had high acceptability by the experts. | AR training may complement traditional methods and useful for novices | Small field of view; discomfort wearing headset; did not visualize needle | N/A |
| Hayasaka et al. (2023) | Measured how well medical students followed textbook needle trajectory for a paramedian epidural in AR, semi-AR, and non-AR conditions. AR improved accuracy, but benefits were mitigated when AR was taken away. | Students using AR rated AR system highly on perspicuity, novelty, stimulation, dependability, and efficiency | Relies on AR marker, which may move during attempted procedure. | Compare AR/MR technology to CT- or ultrasound-guided techniques |
| Huang et al. (2023) | Training with the MR simulator improved participant performance in blind lumbar puncture. | AR/MR useful for learning anatomy and procedure | HoloLens heavy, not friendly to trainees wearing glasses | Develop real-time needle guidance |
| Kim et al. (2023) | Those trained with VR simulation received a higher global rating score and performed the procedure in less time with fewer C-arm uses (compared to those that trained via text and video material). | Participants training using VR reported higher level of satisfaction | Interactions with objects (e.g. needle) in VR does not mimic real life. Dizziness was reported by some participants. | Investigation of VR simulators for various spinal procedures |
| Lau et al. (2023) | Participants strongly agreed that AR software enhances learning and felt it was important to have clear visualization of anatomy and control over how 3D structures are visualized. Integration of haptic devices was desired as a future function. | See previous column | Current system does not allow needle tracking | Link haptic devices and add additional functionality |
| White and Jung (2023) | Perceived subjective improvement and objective improvement noted after 7 minutes of guided instruction using simulator | XR simulation can improve confidence | Cost, calibration issues possible, progressive decline in puncture pad integrity with repeated use | Multicenter study or RCT of high-fidelity neuromodulation training simulation |
| Zheng et al. (2023) | Each intern attempted 30 simulated CSE procedures in a VR environment. Time to successful completion improved, as did global rating score and checklist scores. | 80% of participants felt XR was an excellent substitute to actual procedure | Current system only allows one standardized patient with normal anatomy. Certain steps hard to simulate realistically, including communication with patients. | Compare learning outcomes from VR training to those from clinical practice training |
| ***Preclinical applications of extended reality*** | | | | |
| Ashab et al. (2012) | System provides RMS error 6.6 vs. sonographer measurements. Able to tolerate patient motion up to 10 degrees with 12 mm average error. | Not reported | Requires pre-scan with ultrasound, as well as a separate display. System may give incorrect labels if vertebrae are missed during prescan. | Provide 3D context of underlying anatomy and apply for other procedures |
| Fritz et al. (2012) | AR overlay system accurate, with overall target error 1.9 plus minus 0.9 mm. Virtual epidural spaces successfully entered 20/20 times. | N/A | System prototype costs $13,000, mostly from shielded monitor ~$10000; not tested on live patient, so the impact of motion not tested. | Assess the efficacy of the system for spinal injection procedures in a cadaveric trial and the effectiveness in a clinical trial. |
| Ashab et al. (2013) | Able to identify "lumbar vertebral levels and overlay information onto a live video stream of patient's back" with a mean absolute error ~21% of the vertebral height | Not reported | Same as Ashab et al. (2012) | Additional testing on obese patients |
| Fritz et al. (2013) | All accessible targets (176/187) successfully injected (11 were inaccessible due to anatomy), including 7/7 epidural targets. Median procedure time for epidural injection was 8.6 minutes. No evidence of inadvertent puncture of non-targeted structures such as the dural sac. | N/A | Same as Fritz et al. (2012) | Addition of tilting function to allow non-axial needle paths; plan for clinical trial |
| Hetherington et al. (2017) | "Seventeen of 20 ultrasound scans had successful identification of all vertebral levels" | Not reported | Relies on a specific scan protocol to correctly provide patient-specific data to algorithm. The presented setup uses multiple pieces of additional hardware, but the system is capable of running without said hardware. | Increasing breadth of the training data set to improve accuracy; extend algorithm to whole spine and spinal pathologies (e.g. scoliosis); test system in standard anesthesiology workflow |
| Toews et al. (2018) | Automatic midline detection with system had an RMS error of 2.0 mm when compared to sonographer's manual marking, whereas palpation by anesthesiologist had an RMS error of 5.8. | Not reported | Unable to detect interspinous space | Further investigation in high BMI patients |
| Ameri et al. (2019) | Suggests that AR enhancement of US imaging may reduce rates of inadvertent dural puncture, procedure time, needle path-length to target, and user frustration vs. US alone | Not reported | Displaying 3D imaging on 2D monitor "lacked a sense of depth" | Extend technique to other procedures where needle or a surgical tool is employed |
| Cometa et al. (2020) | Continuous needle advancement during test for LOR produced significantly less needle tip overshoot vs. intermittent needle advancement irrespective of the clinician level of training, technique preference, or LOR depth. | Not reported | Tactile feel of ligamentum flavum not realistic | Verify findings in clinical care |
| Lim et al. (2021) | 8 needle insertions performed per torso phantom. Time and number of x-rays needed were significantly decreased by AR-assistance compared to those of the conventional fluoroscopy-guided approach. Average targeting errors were similar. | Not reported | System relies on multiple pieces of additional equipment, including custom-made 3D pose trackers | N/A |
| Tanwani et al. (2022) | Developed a system that allows holographic visualization of ideal needle trajectory identified using preprocedure ultrasound. 3 or fewer practice repetitions were sufficient for participants to feel comfortable using the system. | Not reported | Does not allow significant patient movement | RCT to evaluate system |
| Jun et al. (2023) | After VR preprocedural planning, needles inserted under AR-assisted vs. conventional methods. AR-assisted insertions required less procedure time and fewer radiographs. No significant targeting errors observed. | Not reported | Requires sophisticated setup with multiple optical trackers and markers | Further studies on patients to understand safety and effectiveness of AR-assisted interventions |
| Reinacher et al. (2023) | First-pass success rates for AR were 82.5% compared to 40% for landmark-based approach; AR allowed better accuracy and faster procedural time | Not reported | Current system cannot account for patient movement; requires preprocedural CT pre-scan in same position as procedure. | Develop dynamic registration of preprocedural images for AR display |
| Wu et al. (2023) | Compared locations of spinous processes as determined using AR to true rotations seen on CT scan.   The maximum mean error along one axis was 4.2 mm. | Not reported | Need for patient to be in same position during CT and AR imaging (used fixation device). Required manual calibration in addition to automatic | Plans RCT to compare success rate of MR-guidance vs. landmark-guided spinal puncture in patients 65+ |
